# Barriers and Facilitators to Equitable Implementation of Long-Acting ART for Adolescents and Youth with HIV in Low- and Middle-Income Settings

**DOI:** 10.1101/2023.11.14.23298511

**Authors:** Nadia A. Sam-Agudu, Chibueze Adirieje, Allison L. Agwu, Natella Rakhmanina

## Abstract

Recent approvals of long-acting (LA) antiretroviral treatment (ART) support an innovative alternative to daily oral pills that can improve adherence and treatment outcomes among adolescents and youth (AY) with HIV. We solicited stakeholder feedback on the implementation of LA ART for AY in low-and middle-income countries (LMICs) through a consensus-building forum at the 2022 International Workshop on HIV and Adolescence.

We used the nominal group technique to generate, record, discuss, vote on, and rank perceived barriers and facilitators to implementing LA ART for AY. All in-person attendees were invited to participate and were assigned to six groups, each representing an intentional mix of AY, clinicians, researchers, program implementers and policymakers. We collected self-reported de-identified demographics and group rankings of barriers and facilitators. Responses were coded and categorized using the social-ecological model’s five levels of influence.

137 Workshop delegates (67.9% male, 27.7% female; 0.7% non-binary and 46.7% less than 35 years old) participated in the group discussions. A large proportion of participants (51.9%) reported working in public health/program implementation. Most participants (88.4%) were from and/or worked in the African region. We identified 55 barriers and 48 facilitators of LA ART implementation and ranked them in social-ecological model categories of public policy, community, institutional/organizational, interpersonal, and individual levels. The highest number of ranked barriers was at the institutional/organizational level. The themes of “equitable access” and “choices of ART” were cross-cutting across individual and interpersonal levels. Other cross-cutting themes were “cost of LA ART” and “need for funding and sustainability of LA ART programs”. Proposed facilitators addressed identified barriers at each social-ecological level of influence and emphasized peer engagement.

Our nominal groups identified key barriers and proposed facilitators at five different social ecological levels, which can inform implementation science-guided design and equitable implementation of youth centered LA ART in LMICs and globally.

## INTRODUCTION

The last decade has seen an improvement in the trend of adolescent AIDS-related mortality [1]. However, there continues to be a disproportionately high rate of adolescent AIDS-related deaths in high-burden African countries (9.92 per 100,000) where ∼80% of adolescents with HIV reside, *vs* globally (2.3 per 100,000) [2]. This disproportionately high HIV-related morbidity and mortality has been ascribed to inadequate access to treatment, low treatment adherence and health services poorly responsive to adolescent needs in Africa [3]. Adolescents and youth (AY) with HIV face many challenges in adherence to daily oral antiretroviral treatment (ART) and have some of the worst treatment outcomes compared to other age groups of people with HIV [4, 5]. To achieve the UNAIDS 95-95-95 goals, adolescents need access to a variety of ART modalities in addition to adherence and retention interventions delivered via innovative, adolescent-responsive strategies [6-9].

Recently approved long-acting (LA) ART for HIV treatment in adolescents and adults represent an attractive alternative to daily oral pills in HIV treatment. These approvals open up opportunities for an innovative biomedical intervention to improve ART adherence and treatment outcomes among AY in low-and middle-income countries (LMICs) [10]. Current options for LA ART comprise several therapeutic choices: a full regimen combination of an integrase strand transfer inhibitor cabotegravir (CAB) and a non-nucleoside reverse transcriptase inhibitor rilpivirine (RPV) (administered as two intramuscular injections every one to two months with optional oral lead in) [11], monoclonal antibody ibalizumab-uiyk (administered as an intravenous infusion or push every two weeks) [12], and a capsid inhibitor lenacapavir (administered as a subcutaneous injection every six months) [13] to be used in combination with other antiretroviral drugs. A study of 303 AY age 13-24 years showed high AY interest in LA ART, with highest interest among those with elevated viral loads [14]. Based on interim data from the IMPAACT MOCHA adolescent study (ClinicalTrials.gov #NCT03497676), LA CAB/RPV ART has been approved by the United States (US) Food and Drug Administration since March 2022 for use among virologically suppressed adolescents and adults ≥12 years old weighing ≥35 kg and without CAB/RPV drug resistance [11]. Additional data on safety, acceptability, tolerability, and pharmacokinetics are anticipated for adolescents 12-17 years from MOCHA in the US, South Africa, Botswana, Puerto Rico, Uganda, and Thailand. Phase III studies among adults with HIV in the USA, Spain, Australia and in seven high-burden African countries demonstrated sustained viral suppression and high acceptance of LA CAB/RPV ART [15-19].

Given its injectable mode of administration and favorable dosing schedule, LA CAB/RPV could become an important option for sustained ART among AY in LMICs [20-25]. Early implementation experience in high-income countries [26-29] can inform some of the programming in LMICs, however, timely stakeholder engagement in LMICs is crucially important to achieve global success in introducing and scaling up this new treatment modality, especially among AY.

A few studies emerging from LMICs are evaluating preferences for, and acceptability of future LA ART among adolescents and youth [30], heralding the timeliness of engaging adolescents and other stakeholders in decision-making regarding this new treatment modality. We aimed to elicit facilitators and barriers to the implementation of LA ART for adolescents and young people with HIV in LMIC settings through a consensus-building forum conducted at the 2022 International Workshop on HIV and Adolescence.

## METHODS

This consensus-building forum was conducted during the in-person International Workshop on HIV and Adolescence held in Cape Town, South Africa on 5^th^ and 6^th^ October 2022 [31]. To address our objective, we adopted the nominal group technique, a structured small group discussion that aims to reach consensus on an issue among stakeholders [32, 33]. This technique combines both qualitative and quantitative methods and is designed to minimize domination of the discussion by individual participants, for example those with more social, economic, or academic power. The nominal group process has four main steps: generate ideas, record the ideas, discuss the ideas, and vote on the ideas to rank them [32] (Fig. 1). A moderator poses a question or questions to participants, who are then asked to prioritize responses given by all group members using numerical rankings. The group needs to come to a consensus on the ranked responses before the responses are finalized. All rapporteurs, selected by the groups, were invited to present a summary of their group’s final consensus on the discussion questions to all Workshop attendees in a plenary session. The nominal group discussions were held during a 60-minute session on Day 2 of the Workshop.

**Figure 1.**
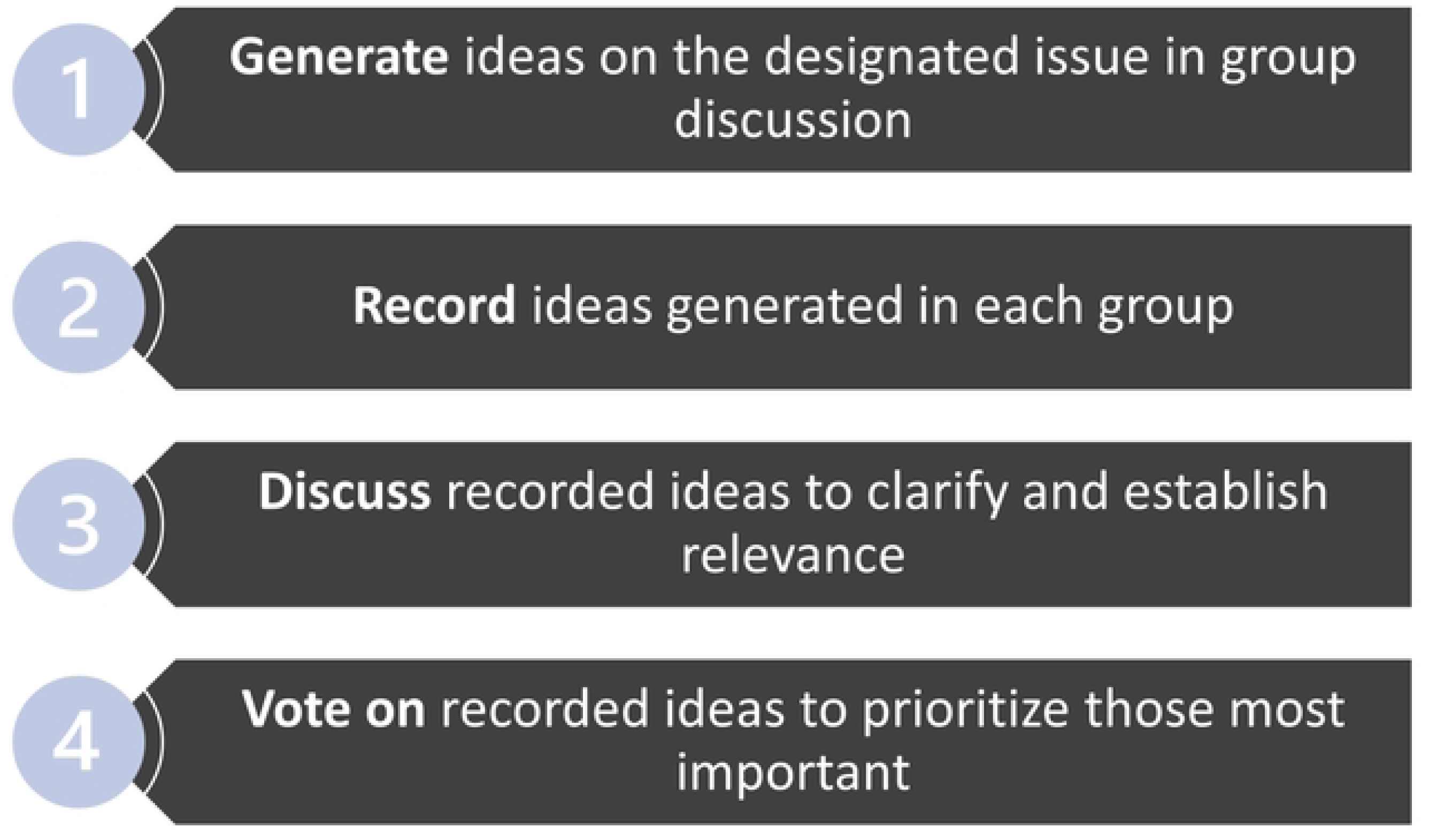
Process of the nominal group technique for reaching consensus among stakeholders.

### Workshop Delegate Recruitment

A total of 263 registered delegates attended the Workshop, all of whom were invited to participate in the forum. Prior to the nominal group discussions, there were three brief presentations totaling 20 minutes: first presentation by author AA reviewed experiences and data from the US on adolescent preferences for LA ART; NR described practical experiences in administering LA ART to adolescents and young adults with HIV; and NASA explained the nominal group process to all attendees, informing them of the intent to write a report and requesting their voluntary participation. Workshop delegates who proceeded to participate in the group discussion after this point were considered to have provided consent. After these three presentations, an opportunity was given for attendees to ask any clarifying questions. Of note, there were two plenary Day 1 presentations on HIV prevention and treatment for AY which had included information on LA antiretrovirals [31].

### Data Collection

Workshop delegates were assigned to six groups, each representing an intentionally diverse mix of youth, clinicians, researchers, program implementers and policymakers. Group assignments were based on participants’ self-identification of their role or job description during Workshop registration. Two moderators were assigned per group: a youth implementer or advocate and a senior researcher, implementer, or clinician with expertise in adolescent HIV. Each group nominated a rapporteur to document group member responses and rankings for questions posed by the moderators. Group discussions were conducted for approximately 40 minutes. A standard form was used to collect self-reported de-identified demographic data from each participant, including region and country of work and/or residence, gender, age range, and area of focus for their work. Participants were instructed (verbally and in writing) not to write any identifying information on their individual forms. Moderators used a second, separate standard form to document group responses and rankings related to the study questions. There were no pictures taken, nor video or audio recordings; all data collected were exclusively hand-written on standard hard-copy forms and subsequently transferred to an Excel sheet for analysis.

The key questions posed to participants for group discussion were:

- *What are the barriers to implementing (rolling out) long-acting ART for adolescents and young adults in your country or region?* Write down all barriers, then rank the top three.
- *What are the facilitators, or potential solutions in addressing gaps to implement (roll out) long-acting ART for adolescents and young adults in your country or region?* Write down all the facilitators/potential solutions, then rank the top three.

### Data Analysis

Demographic data were analyzed by descriptive statistics. Nominal group technique responses and rankings regarding barriers and facilitators to the implementation of LA ART were coded and categorized using the social-ecological model. This model considers the different levels of influence on the adoption, uptake or implementation of a health behavior and/or intervention [34]. The five levels of influence applied to this analysis were: individual (regarding the adolescent with HIV); interpersonal (family, friends, and social networks of the adolescent); institutional/organizational (health facilities, schools, religious centers, workplaces); community (culture, informal or formal social norms, and community as a place of residence); and health-related policy (national, state, and other local laws). Group responses were also coded and categorized according to cross-cutting issues at two or more of the social-ecological levels. The four-member analysis team (NASA, CA, ALA and NR) coded and categorized the nominal group technique data independently; individual analyses were then merged and discussed as a team. Any conflicting or outlying areas of analysis were voted on for consensus.

### Ethical Considerations

Ethical oversight was waived by the University of Maryland Baltimore, given that participants were discussing questions posed at a public forum during a scientific workshop and that demographic data were voluntarily self-reported without any personal identifiers. Delegates had the option not to participate if they did not wish to do so. As this session was listed as part of the conference agenda that was published in advance, any minors under 18 years were to be included for participation if they had registered for and were attending the workshop in person. The findings of this forum are being disseminated as a conference report.

## RESULTS

A total of 137 Workshop delegates (52.1% of the 263 in-person attendees) participated in the nominal group discussions, of whom 93 (67.9%) self-reported as male, 38 (27.7%) as female, and 1 (0.7%) as non-binary (Table 1). With respect to age, 11 (8.0%) participants reported being 20-24 years old, 53 (38.7%) were 25-34 years old, and 68 (49.6%) were ≥35 years old; there were no participants <20 years old. In terms of profession or roles, the largest proportion of participants (49.6%) reported working in public health or program implementation.

**Table 1.** Demographic Profile of Participants in Nominal Groups, N=137.

| Characteristic | n | % |
| --- | --- | --- |
| **Gender** |  |  |
| Male | 93 | 67.9 |
| Female | 38 | 27.7 |
| Transgender/Non-binary | 1 | 0.7 |
| Undisclosed | 5 | 3.6 |
| **Age Group (years)** |  |  |
| 10-19 | 0 | 0.0 |
| 20-24 | 11 | 8.0 |
| 25-34 | 53 | 38.7 |
| $\geq 35$ | 68 | 49.6 |
| Undisclosed | 5 | 3.6 |
| **Profession/Practice** |  |  |
| Public Health/Program Implementation<sup>a</sup> | 68 | 49.6 |
| Research<sup>a</sup> | 49 | 35.8 |
| Public Health/Program Implementation<br>and Research | 6 | 4.4 |
| Public Health/Program Implementation<br>and Policy | 2 | 1.5 |
| Research and Policy | 1 | 0.7 |
| Policy only | 3 | 2.2 |
| Activist<sup>b</sup> | 11 | 8.0 |
| Other<sup>c</sup> | 7 | 5.1 |
| Undisclosed | 6 | 4.4 |

<sup>a</sup> Includes all participants who indicated this exclusively or in combination with another occupation (may add up to >100%)
<sup>b</sup> Includes 1 participant engaged in both Activism and Research
<sup>c</sup> Includes Advocacy (1), Clinician (1), Clinical psychologist (1), Industry (2), Pharmaceutical (1), and Marketing (1)

In terms of region of primary residence and/or work, most participants (88.4%) were from, and/or worked in the African region, followed by North America (8.2%) and Europe (1.4%) (Figure 2). Four percent of all participants reported living and working in both Africa and North America.

**Figure 2.**
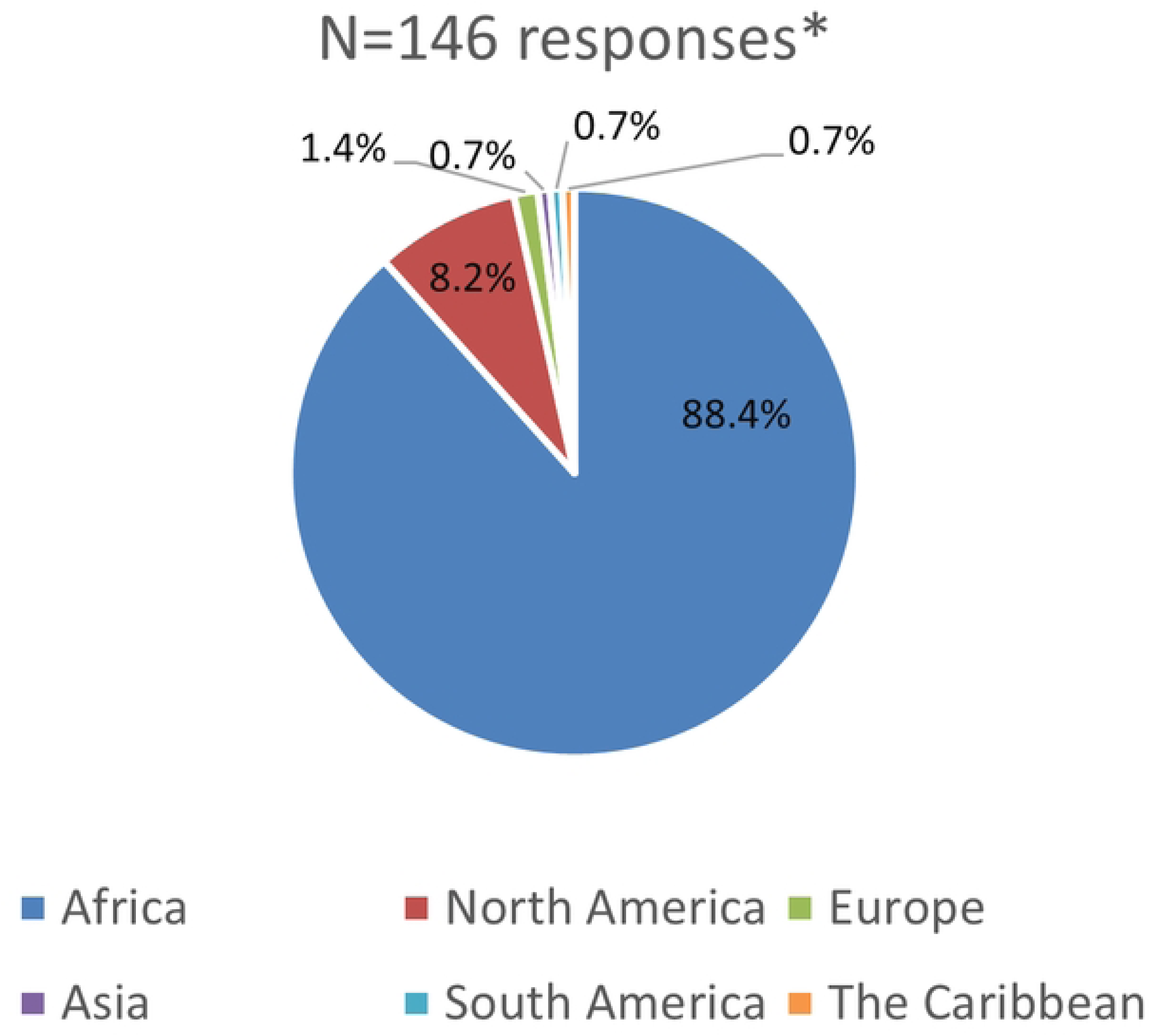
Region of primary residence and/or work among participants based on responses. \**Pie chart shows regional representation at the Workshop; some participants lived or worked in more than one region, with each recorded separately. There were no participants from Oceania*.

At country level, South Africa had the most representation (57%) among all participants, which is expected given that the conference was held in Cape Town. Eswatini (6.3%), Namibia (5.5%), Zimbabwe (4.7%), Kenya (3.9%) and Uganda (3.9%) rounded up the top six African countries by participant representation (Figure 3). Among the six non-African countries reported, the USA was represented by nine (60%) of 15 participants, two for the United Kingdom (13.3%), and one each (6.7%) for Haiti, Malaysia, France, and Switzerland (not displayed).

**Figure 3.**
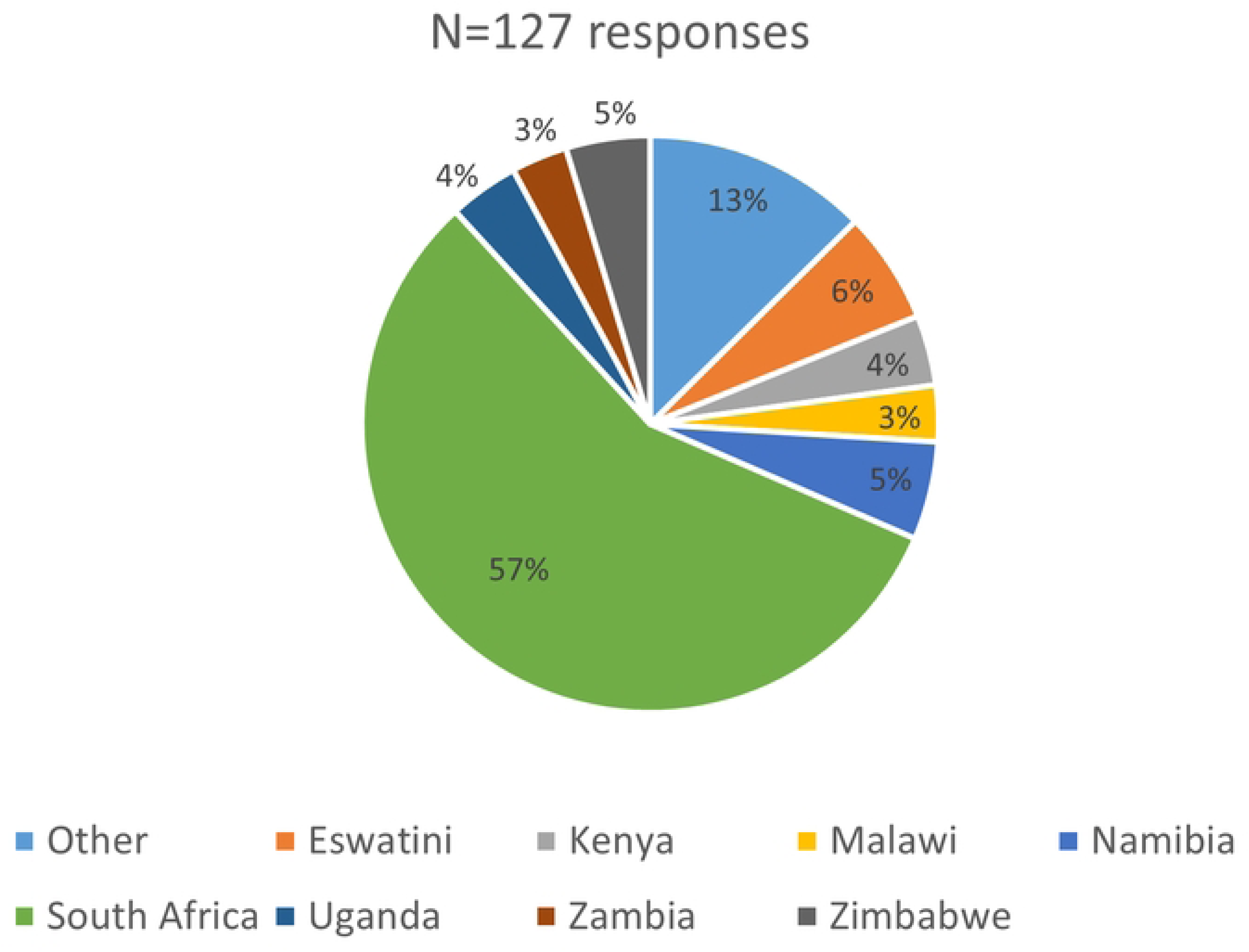
Country of primary residence and/or work among participants living and/or working in Africa. *“Other” refers to countries that each represent 1-2% of responses provided (Botswana, Burundi, Cameroon, DR Congo, Ethiopia, Mozambique, Rwanda, Senegal, Somalia and Tanzania)*.

### Consensus on LA ART for Adolescents and Youth

A total of 55 barriers and 48 facilitators relevant to LA ART implementation were provided by the groups (Supporting Information, Appendix 1). The data were collated, and then merged to harmonize similar concepts. All barriers and facilitators were then sorted according to the five levels of the social-ecological model, in addition to a cross-cutting category.

The top three ranked barriers and facilitators from each group were extracted, collated, and sorted according to the social-ecological model (Figure 4). These collated factors were listed in order of priority, according to the number of times they were listed in individual group rankings across the six groups.

**Figure 4.**
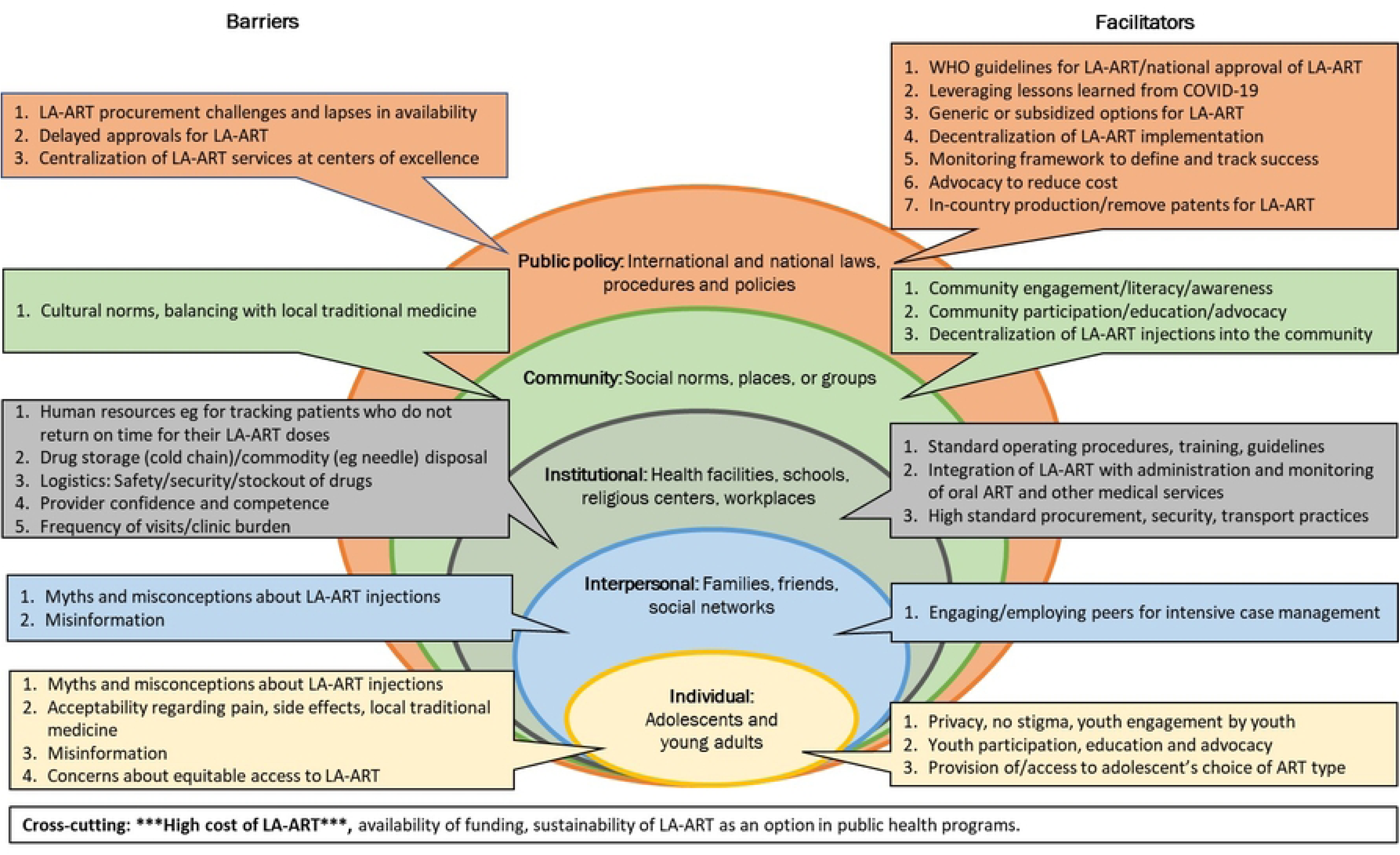
Top-ranked barriers and facilitators to implementing LA ART for adolescents and youth. *There are less than or more than three factors at some social-ecological barrier or facilitator levels, because between zero and 18 responses (the top three for each of the six groups) were ranked. To minimize analytical bias, group responses were not requested by social ecological level. “Cost of LA ART/procurement challenges” was listed in the top three barriers for all six groups.*

The highest ranked *public policy* barriers included delayed approval, and challenges with procurement, availability, and centralized distribution of LA ART on a national level. To mitigate these barriers, the groups prioritized several strategies including clear national and World Health Organization (WHO) recommendations on the use of LA ART, patent-free in-country production, use of generic or subsidized reduced-cost CAB/RPV, and decentralization of LA ART procurement and delivery services. Further supportive public policy strategies proposed included close monitoring and evaluation of treatment outcomes and applying lessons learned from testing and immunization campaigns in the COVID-19 pandemic response.

Prominent barriers at the *community level* included negative cultural norms and beliefs around injections, and the use/promotion of local traditional medicines instead of ART for HIV treatment. To address this challenge, groups highlighted the need for early community awareness and engagement, education and support, participation and advocacy, and importantly, decentralization of LA ART within the community.

The highest number of ranked barriers was at the *institutional/organizational* level (Fig. 4) and included a lack of confident and competent personnel to administer injections and track and support AY on ART. In addition, the complex logistics of drug storage and delivery and expected increased clinic visit burden were cited as barriers affecting healthcare providers and facilities. The proposed solutions included trainings, development of standard operating procedures, integration of LA ART within oral ART programs and other medical services and optimized secured procurement and transportation of the LA drugs.

Misinformation, misconceptions, and myths about LA ART were prominent barriers at the *interpersonal* and *individual* levels. Participants emphasized youth participation and advocacy, and engaging peer advocates for service delivery using intensive case management models. The themes of “equitable access” and “choices of ART” were cross-cutting in the individual and interpersonal levels for all six groups. Other cross-cutting themes were high cost of LA ART, and the need for funding and sustainability of LA ART programs once established.

## DISCUSSION

We report consensus findings on the implementation of LA ART among adolescents and youth with HIV from a large group of stakeholders (nearly 50% less than 35 years old) attending an international workshop on HIV in young people. A major strength of this report is that participants include youth advocates and multidisciplinary frontline programmatic and research staff working with these populations across multiple LMICs, including in Africa where the HIV epidemic is disproportionately occurring. Our nominal group technique and analysis identified key barriers at five different social ecological levels. We synthesized these barriers and the proposed tangible solutions and strategies for consideration when designing youth centered LA ART programs, especially those relevant for African countries.

Along the five social ecological levels, several recurrent themes emerged relating to barriers and facilitators. Not surprisingly and in accordance with published multi-country analysis on considerations for the rollout of novel ART [35], the highest number of ranked barriers in our analysis was at the institutional/organizational level. These barriers included the need for a competent workforce and confident healthcare workers, increased number patients at clinic visits, changes in clinic flow and patient follow-up protocols, and multiple logistics of cold-chain storage of LA CAB/RPV and disposal of supplies. These barriers align and resonate with the body of evidence on the ongoing need for multi-level health system strengthening and investment into human resources [36]. Based on participants’ feedback and suggested pathways for LA ART implementation, we argue that integration of LA ART into existing ART programs provides a unique opportunity to update and optimize HIV-related trainings, standard operating procedures, patient outcomes monitoring and intensified focus on quality of care in clinical settings [36, 37].

Reminiscent of recent experiences with COVID-19 vaccines [38, 39], misinformation and misconceptions about LA ART within communities and among AY were perceived as prominent barriers at interpersonal and individual levels. Lack of attention to interpersonal and social dynamics limits our understanding of the effective strategies needed to expand and scale up ART access, for example in high HIV burden African countries [40]. As suggested by Myburgh et al., it is not enough to place predominant focus on health system infrastructure when expanding ART access; innovative individual and interpersonal-level strategies are additionally required to “increase the number of clients on ART while sustaining current processes to retain all in care” [40]. Based on the individual and interpersonal level solutions provided by our participants, providing transparent and accurate information, addressing myths and misperceptions, optimizing peer education, engagement, and advocacy, and reducing/eliminating stigma will all be crucially important for successful LA ART roll out for AY. These barrier-focused solutions are consistent with the principles of differentiated service delivery and high quality of care in HIV programs as described by the WHO [37, 41].

The public policy barriers and solutions described highlight the need for clear global and national guidance on the use of LA ART and facilitated access to patent-free generic or subsidized LA CAB/RPV formulations through decentralized mechanisms. These solutions depend on affordability of LA ART formulations and equitable access in LMICs, similar to what has been negotiated and achieved for LA CAB pre-exposure prophylaxis (PrEP) [42, 43]. As with LA CAB PrEP [44], cost should not be allowed to derail the scale up of new ART modalities and their availability to eligible patients, including AY, for whom they hold promise to improve adherence and treatment outcomes [21]

Our consensus findings provide valuable information for implementation science-guided roll-out of LA ART for AY in LMICs. Participants identified and ranked barriers and facilitators across individual, interpersonal, institutional, community and public policy domains. The results and analysis are extremely useful for identifying, selecting and developing highly relevant implementation strategies to minimize barriers and enhance facilitators across multiple social-ecological levels. In addressing the determinants (barriers and facilitators) of successful implementation, implementation strategies facilitate desired implementation outcomes, which, for AY-targeted LA ART, would include Acceptability among AY and other key stakeholders, Uptake among AY, Adoption by healthcare systems and providers, Feasibility, Fidelity, Costs, and Sustainment/Sustainability [45, 46].

Our report is not without limitations. For a forum on adolescent HIV, there were no participants aged 10-19 years: <10% were 20-24 years old, and less than a third of participants were female. Adolescent girls and young women carry a disproportionally high burden of HIV in African countries and LMICs and bear a significant responsibility in preventing perinatal HIV transmission [47]; they were, unfortunately, under-represented in this forum. However, overall, nearly half of our participants were aged <35 years. Furthermore, we assigned youth moderators and participants to each group to amplify the voices of AY participants. Despite these limitations, to date, this is the largest stakeholder consultation on youth focused implementation of LA ART in LMICs that has been conducted shortly after the approval and roll out of this ART modality for AY in resource-rich settings.

### Conclusions

LA ART presents an attractive alternative to daily oral treatment for HIV, especially for adolescents and youth with adherence issues. Our stakeholder consensus findings come at a time when most LMICs, including high burden African countries, are still at the pre-or early implementation stage of LA ART for any population; even high-income countries with LA ART approval are reporting implementation challenges with logistics and uptake [48]. Guided by lessons learned from consultations such as we have described, implementation of LA ART for AY will allow HIV programs to enhance community engagement, strengthen AY focused health services, negotiate affordable costs, and invest in quality and sustainable logistics and supply chains. We now have a window of opportunity for implementation science-guided approaches and early and meaningful engagement with AY and other key stakeholders to achieve equitable LA ART implementation for current and future generations.

## Data Availability

All of the data collected is provided in the manuscript and supporting materials.

## ACKNOWLEDGEMENTS

The authors would like to thank Academic Medical Education and the Organizing and Scientific Committees of the 2022 International Workshop on HIV and Adolescence for their enabling support. We extend our appreciation to the moderators and to the Workshop delegates who participated in the nominal group sessions and shared their opinions and experiences.

## SUPPORTING INFORMATION

***S1: List of Barriers and Facilitators Identified by the Nominal Groups.*** Database of 55 barriers and 48 facilitators relating to roll-out of long-acting ART that was provided by all six nominal groups.

## REFERENCES

1. Slogrove AL, Sohn AH. The global epidemiology of adolescents living with HIV: time for more granular data to improve adolescent health outcomes. Curr Opin HIV AIDS. 2018;13(3):170–8.

2. UNICEF. UNICEF Data: Adolescent HIV Treatment 2021 [Available from: https://data.unicef.org/topic/hivaids/adolescent-hiv-treatment/.

3. Ammon N, Mason S, Corkery JM. Factors impacting antiretroviral therapy adherence among human immunodeficiency virus-positive adolescents in Sub-Saharan Africa: a systematic review. Public health. 2018;157:20–31.

4. Frescura L, Godfrey-Faussett P, Feizzadeh AA, El-Sadr W, Syarif O, Ghys PD. Achieving the 95 95 95 targets for all: A pathway to ending AIDS. PloS one. 2022;17(8):e0272405.

5. Zhou S, Cluver L, Shenderovich Y, Toska E. Uncovering ART adherence inconsistencies: An assessment of sustained adherence among adolescents in South Africa. Journal of the International AIDS Society. 2021;24(10):e25832.

6. Daltro ACB, Almeida CS, Unfried AGC, de Aquino TR, Travassos A. Virological failure and adherence to antiretroviral therapy in adolescents and young adults living with human immunodeficiency virus. Tropical medicine & international health : TM & IH. 2023;28(3):162–74.

7. Ridgeway K, Dulli LS, Murray KR, Silverstein H, Dal Santo L, Olsen P, et al. Interventions to improve antiretroviral therapy adherence among adolescents in low-and middle-income countries: A systematic review of the literature. PloS one. 2018;13(1):e0189770.

8. Chem ED, Ferry A, Seeley J, Weiss HA, Simms V. Health-related needs reported by adolescents living with HIV and receiving antiretroviral therapy in sub-Saharan Africa: a systematic literature review. Journal of the International AIDS Society. 2022;25(8):e25921.

9. UNAIDS. Understanding Fast Track: Accelerating Action to End the AIDS Epidemic by 2030 2015 [Available from: https://www.unaids.org/sites/default/files/media_asset/201506_JC2743_Understanding_FastTrack_en.pdf.

10. Venkatesan P. Long-acting injectable ART for HIV: a (cautious) step forward. Lancet Microbe. 2022;3(2):e94.

11. US Food and Drug Administration. CABENUVA [package insert] 2022 [Available from: https://www.accessdata.fda.gov/drugsatfda_docs/label/2022/212888s005s006lbl.pdf.

12. US Food and Drug Administration. Trogarzo (ibalizumab-uiyk) [package insert] 2020 [Available from: https://www.accessdata.fda.gov/drugsatfda_docs/label/2020/761065s008lbl.pdf.

13. US Food and Drug Administration. SUNLENCA [package insert] 2022 [Available from: https://www.accessdata.fda.gov/drugsatfda_docs/label/2022/215973s000lbl.pdf.

14. Weld ED, Rana MS, Dallas RH, Camacho-Gonzalez AF, Ryscavage P, Gaur AH, et al. Interest of Youth Living With HIV in Long-Acting Antiretrovirals. Journal of acquired immune deficiency syndromes. 2019;80(2):190–7.

15. Orkin C, Oka S, Philibert P, Brinson C, Bassa A, Gusev D, et al. Long-acting cabotegravir plus rilpivirine for treatment in adults with HIV-1 infection: 96-week results of the randomised, open-label, phase 3 FLAIR study. The lancet HIV. 2021;8(4):e185–e96.

16. Orkin C, Bernal Morell E, Tan DHS, Katner H, Stellbrink HJ, Belonosova E, et al. Initiation of long-acting cabotegravir plus rilpivirine as direct-to-injection or with an oral lead-in in adults with HIV-1 infection: week 124 results of the open-label phase 3 FLAIR study. The lancet HIV. 2021;8(11):e668–e78.

17. Swindells S, Lutz T, Van Zyl L, Porteiro N, Stoll M, Mitha E, et al. Week 96 extension results of a Phase 3 study evaluating long-acting cabotegravir with rilpivirine for HIV-1 treatment. AIDS (London, England). 2022;36(2):185–94.

18. Bares SH, Scarsi KK. A new paradigm for antiretroviral delivery: long-acting cabotegravir and rilpivirine for the treatment and prevention of HIV. Curr Opin HIV AIDS. 2022;17(1):22–31.

19. Taki E, Soleimani F, Asadi A, Ghahramanpour H, Namvar A, Heidary M. Cabotegravir/Rilpivirine: the last FDA-approved drug to treat HIV. Expert Rev Anti Infect Ther. 2022;20(8):1135–47.

20. Freij BJ, Aldrich AM, Ogrin SL, Olivero RM. Long-Acting Antiretroviral Drug Therapy in Adolescents: Current Status and Future Prospects. J Pediatric Infect Dis Soc. 2023;12(1):43–8.

21. Abrams EJ, Capparelli E, Ruel T, Mirochnick M. Potential of Long-Acting Products to Transform the Treatment and Prevention of Human Immunodeficiency Virus (HIV) in Infants, Children, and Adolescents. Clinical infectious diseases : an official publication of the Infectious Diseases Society of America. 2022;75(Suppl 4):S562–S70.

22. Culhane J, Sharma M, Wilson K, Roberts DA, Mugo C, Wamalwa D, et al. Modeling the health impact and cost threshold of long-acting ART for adolescents and young adults in Kenya. EClinicalMedicine. 2020;25:100453.

23. Thoueille P, Choong E, Cavassini M, Buclin T, Decosterd LA. Long-acting antiretrovirals: a new era for the management and prevention of HIV infection. J Antimicrob Chemother. 2022;77(2):290–302.

24. Scarsi KK, Swindells S. The Promise of Improved Adherence With Long-Acting Antiretroviral Therapy: What Are the Data? Journal of the International Association of Providers of AIDS Care. 2021;20:23259582211009011.

25. Kerrigan D, Mantsios A, Gorgolas M, Montes ML, Pulido F, Brinson C, et al. Experiences with long acting injectable ART: A qualitative study among PLHIV participating in a Phase II study of cabotegravir + rilpivirine (LATTE-2) in the United States and Spain. PloS one. 2018;13(1):e0190487.

26. McGowan JP, Fine SM, Vail RM, Merrick ST, Radix AE, Gonzalez CJ, et al. Use of Injectable CAB/RPV LA as Replacement ART in Virally Suppressed Adults. New York State Department of Health AIDS Institute Clinical Guidelines. Baltimore (MD)2022.

27. Rakhmanina N, Richards K, Adeline Koay WL. Transient Viremia in Young Adults With HIV After the Switch to Long-Acting Cabotegravir and Rilpivirine: Considerations for Dosing Schedule and Monitoring. Journal of acquired immune deficiency syndromes. 2023;92(3):e14–e7.

28. Christopoulos KA, Grochowski J, Mayorga-Munoz F, Hickey MD, Imbert E, Szumowski JD, et al. First Demonstration Project of Long-Acting Injectable Antiretroviral Therapy for Persons With and Without Detectable Human Immunodeficiency Virus (HIV) Viremia in an Urban HIV Clinic. Clinical Infectious Diseases. 2022;76(3):e645–e51.

29. Kilcrease C, Yusuf H, Park J, Powell A, Rn LJ, Rn JO, et al. Realizing the promise of long-acting antiretroviral treatment strategies for individuals with HIV and adherence challenges: an illustrative case series. AIDS Res Ther. 2022;19(1):56.

30. Toska E, Zhou S, Chen-Charles J, Gittings L, Operario D, Cluver L. Factors Associated with Preferences for Long-Acting Injectable Antiretroviral Therapy Among Adolescents and Young People Living with HIV in South Africa. AIDS and behavior. 2023;27(7):2163–75.

31. Academic Medical Education. International Workshop on HIV & Adolescence 2022 2022 [Available from: https://academicmedicaleducation.com/meeting/hiv-adolescence-2022.

32. US Centers for Disease Control and Prevention. Gaining Consensus Among Stakeholders Through the Nominal Group Technique 2018 [Available from: https://www.cdc.gov/healthyyouth/evaluation/pdf/brief7.pdf.

33. Van de Ven AH, Delbecq AL. The nominal group as a research instrument for exploratory health studies. American journal of public health. 1972;62(3):337–42.

34. US Centers for Disease Control and Prevention. Models and Frameworks for the Practice of Community Engagement. Principles of Community Engagement. 2nd Ed. ed2015.

35. Mantsios A, Murray M, Karver TS, Davis W, Galai N, Kumar P, et al. Multi-level considerations for optimal implementation of long-acting injectable antiretroviral therapy to treat people living with HIV: perspectives of health care providers participating in phase 3 trials. BMC health services research. 2021;21(1):255.

36. Mikkelsen E, Hontelez JA, Jansen MP, Barnighausen T, Hauck K, Johansson KA, et al. Evidence for scaling up HIV treatment in sub-Saharan Africa: A call for incorporating health system constraints. PLoS medicine. 2017;14(2):e1002240.

37. World Health Organization. Maintaining and improving quality of care within HIV clinical services. 2019 [Available from: https://apps.who.int/iris/handle/10665/325857.

38. Nachega JB, Sam-Agudu NA, Masekela R, van der Zalm MM, Nsanzimana S, Condo J, et al. Addressing challenges to rolling out COVID-19 vaccines in African countries. Lancet Glob Health. 2021;9(6):e746–e8.

39. Moola S, Gudi N, Nambiar D, Dumka N, Ahmed T, Sonawane IR, et al. A rapid review of evidence on the determinants of and strategies for COVID-19 vaccine acceptance in low-and middle-income countries. J Glob Health. 2021;11:05027.

40. Myburgh H, Reynolds L, Hoddinott G, van Aswegen D, Grobbelaar N, Gunst C, et al. Implementing ’universal’ access to antiretroviral treatment in South Africa: a scoping review on research priorities. Health Policy Plan. 2021;36(6):923–38.

41. Grimsrud A, Walker D, Ameyan W, Brusamento S. Providing differentiated delivery to children and adolescents. WHO Policy Brief, [Internet]. 2019 July 11, 2023. Available from: https://www.who.int/publications/i/item/WHO-CDS-HIV-19.25.

42. World Health Organization. WHO applauds agreement to scale-up generic manufacturing for access to long-acting injectable antiretrovirals 2022 [Available from: https://www.who.int/news/item/28-07-2022-who-applauds-agreement-to-scale-up-generic--manufacturing-for-access-to-long-acting-injectable-antiretrovirals.

43. World Health Organization. Guidelines on long-acting injectable cabotegravir for HIV prevention 2022 [Available from: https://www.who.int/publications/i/item/9789240054097.

44. The Lancet H. Equitable access to long-acting PrEP on the way? The lancet HIV. 2022;9(7):e449.

45. Vorkoper S, Sam-Agudu NA, Bekker LG, Sturke R. Implementation Science for Eliminating HIV Among Adolescents in High-Burden African Countries: Findings and Lessons Learned from the Adolescent HIV Prevention and Treatment Implementation Science Alliance (AHISA). AIDS and behavior. 2023;27(Suppl 1):3–6.

46. Aarons GA, Reeder K, Sam-Agudu NA, Vorkoper S, Sturke R. Implementation determinants and mechanisms for the prevention and treatment of adolescent HIV in sub-Saharan Africa: concept mapping of the NIH Fogarty International Center Adolescent HIV Implementation Science Alliance (AHISA) initiative. Implement Sci Commun. 2021;2(1):53.

47. UNICEF. UNICEF Data: Adolescent HIV Treatment 2022 [Available from: https://data.unicef.org/topic/hivaids/adolescent-hiv-treatment/.

48. Collins LF, Corbin-Johnson D, Asrat M, Morton ZP, Dance K, Condra A, et al. Early Experience Implementing Long-Acting Injectable Cabotegravir/Rilpivirine for Human Immunodeficiency Virus-1 Treatment at a Ryan White-Funded Clinic in the US South. Open Forum Infect Dis. 2022;9(9):ofac455.

